# Causal linkage of tobacco smoking with ageing traits: a Mendelian randomization analysis towards telomere attrition and frailty

**DOI:** 10.1101/2022.06.27.22276928

**Authors:** Sehoon Park, Seong Geun Kim, Soojin Lee, Yaerim Kim, Semin Cho, Kwangsoo Kim, Yong Chul Kim, Seung Seok Han, Hajeong Lee, Jung Pyo Lee, Kwon Wook Joo, Chun Soo Lim, Yon Su Kim, Dong Ki Kim

## Abstract

**Background:** Ageing traits and frailty are important health issues in modern medicine. Evidence supporting the causal effects from tobacco smoking on various ageing traits is warranted.

**Methods:** This Mendelian randomization (MR) analysis instrumented 377 genetic variants associated with being an ever smoker in a genome-wide significant level to test the causal estimates from tobacco smoking. The outcome data included 337,318 white British ancestry UK Biobank participants. Leukocyte telomere length, appendicular lean mass index, subjective walking pace, handgrip strength, and wristband accelerometry determined physical activity degree were collected as ageing related outcomes. Summary-level MR by inverse variance weighted method and pleiotropy-robust MR methods, including weighted median and MR–Egger, was performed.

**Results:** Summary-level MR analysis indicated that higher genetic predisposition for tobacco smoking was significantly associated with shorter leukocyte telomere length [2-fold prevalence increase in smoking towards standardized Z-score, -0.041 (-0.054, -0.028)], lower appendicular lean mass index [-0.007 (-0.010, -0.005)], slower walking pace [ordinal category, -0.047 (-0.054, -0.033)], and lower time spent on moderate-to-vigorous physical activity [hours per week, -0.39 (-0.56, -0.23). The causal estimates were nonsignificant towards handgrip strength phenotype [kg, 0.074 (-0.055, 0.204)]. Pleiotropy-robust MR results generally supported the main causal estimates.

**Conclusion:** Genetically predicted tobacco smoking is significantly associated with worse ageing phenotypes. Healthcare providers may continue to reduce tobacco use which may be helpful to reduce the burden related to ageing and frailty.

## Background

Ageing population epidemic is an important socioeconomic burden.^1,2^ Global population aged 60 and over is above one billion, and the number is rapidly increasing. Ageing is an important health status related to frailty syndrome,^3^ a state with muscle loss and low physical activity. Ageing occurs differently among individuals and those with accelerated ageing has higher risk of cardiovascular disease and mortality.^4-6^

Tobacco smoking is another important global health problem which increases risks of cardiovascular disease, malignancy, and death.^7^ The causal linkage between tobacco smoking and ageing traits needs further study. Tobacco exposure causes cellular senescence,^8^ thus, it is highly assumed that biological ageing would be accelerated by tobacco use. Despite some observational studies supporting the association,^9^ the direct causal effect is difficult to be revealed by conventional study design because of complex confounding and reverse causal effects.^10^ A higher level evidence suggesting the causal effect would be important, as it would firmly support that social interventions to reduce tobacco use would be helpful to reduce the rapidly increasing burden from ageing.

In recent biobank databases (e.g. UK Biobank), ageing related traits including leukocyte telomere length (indicator for biologic ageing), sarcopenia phenotypes (appendicular lean mass index, walking pace, and handgrip strength), and objectively measured physical activity traits are available in population-scale.^11^ The availability of the large-scale ageing traits with associated genotypes enabled various studies studying the risk factors and outcomes related to ageing. Particularly, genetic epidemiology analysis, Mendelian randomization (MR), has been widely performed. MR utilizes genetic instrumental variable, which is minimally biased by confounding effects or reverse causation as it is fixed before birth, to demonstrate causal effect.^10^ MR analysis has been performed also in studies related to ageing phenotypes and the method would be a useful tool to investigate the causal effects from tobacco smoking on ageing.^12-14^

In this study, we hypothesized that the causal effects from tobacco smoking on various ageing traits would be present. We performed MR analysis in the UK Biobank implementing genetic instruments developed from a large-scale genome-wide association study (GWAS) meta-analysis towards tobacco smoking phenotype. We found tobacco smoking may causally affect accelerating biologic ageing, physical inactivity, and sarcopenia.

## Methods

### Ethical considerations

The study was approved by the institutional review boards of Seoul National University Hospital (No. E-2203-053-1303) and the UK Biobank consortium (application No. 53799).^11,15^ The study was performed in accordance with the Declaration of Helsinki. The requirement for informed consent was waived by the attending institutional review boards because this study investigated public anonymous database without medical intervention.

### Genetic instruments for tobacco smoking

We used the genetic variants for tobacco smoking from one of the largest GWAS meta-analysis for 1.2 million European ancestry individuals including over half a million ever smokers.^16^ There were 29 studies included in the meta-analysis and the 23andMe, UK Biobank, deCODE, and HUNT were the representative studies which provided relatively large number of samples. The 378 lead single nucleotide polymorphisms (SNPs) towards the phenotype of being ever a regular smoker (smoking initiation) with conditionally genome-wide significant association were selected as genetic instruments for tobacco smoking (Supplementary Table 2).

Genetic instruments for MR analysis should attain three core assumptions to demonstrate causal estimates.^10^ First, the relevance assumption is that the instruments should be closely associated with the exposure phenotype, and this was considered attained as the SNPs were identified from the large-scale GWAS meta-analysis. Moreover, the genome-wide significant SNPs explained ∼2% variance for smoking initiation, providing sufficient power for MR analysis considering the size of the outcome dataset.^16^ Second, the independence assumption is that the instruments should not be associated with confounding phenotypes. We used pleiotropy-robust MR analysis methods (MR-Egger, weighted median) which relaxes the assumption to demonstrate that the causal estimates are not affected from such pleiotropy.^17,18^ In addition, MR-Egger intercept P value was calculated to test the likelihood of presence of a directional pleiotropy. Furthermore, we performed sensitivity analysis with additional exclusion for the SNPs genome-wide significantly associated with other phenotypes in the GWAS catalog, which the search was already performed in the previous GWAS meta-analysis.^16,19^ Third, the exclusion-restriction assumption is that the causal pathway should be through the exposure of interest. We used the weighted median method which relaxes the assumption for up to half of instrumented genetic weights.^18^ In addition, a direct effect from a SNP towards an outcome phenotype would invalidate the assumption, thus, we disregarded SNPs with direct genome-wide significant association (P < 5E-8) with an outcome phenotype when performing the analysis.

### UK Biobank outcome data

UK Biobank data is a prospective cohort of ∼ 500,000 individuals aged 50-69 years old at baseline.^11,15^ The database includes deep phenotyping and genotyping information for the samples, and has been widely used in modern medical researches. Because of the large-sample size, availability for diverse phenotypes, and large-scale genotype data, UK Biobank data has been used in various MR studies.^12-14,20^

We used the data from white British ancestry samples in the UK Biobank data, as ethnic-specific analysis is important for MR analysis and the genetic instruments were developed from European ancestry population.^19,20^ After disregarding those with excess kinship, sex chromosomal aneuploidy, or outliers regarding heterogeneity or missing rate, total of 337,318 white British ancestry individuals were included for the analysis.

### Outcome ageing traits

We collected diverse traits related to ageing or frailty. 1) First, leukocyte telomere length was collected.^13^ The phenotype was used to reflect proxies of biological ageing, as those with accelerated ageing exhibits shortening of telomere. The log-transformed and standardized Z scores for leukocyte telomere length was collected in 326,075 samples. 2) Second, we collected an objective parameter for muscle mass loss, the appendicular lean mass adjusted for body mass index (appendicular lean mass/body mass index), measured by a BC418MA body composition analyzer (Tanita, Tokyo, Japan) and available in 329903 samples.^12,21^ 3) Third, we collected functional parameter for sarcopenia including walking pace, self-reported as slow, average, or brisk category, and handgrip strength, measured isometrically using a Jamar J00105 hydraulic hand dynamometer (Lafayette Instrument Company, IN, USA).^12^ The information for walking pace was collected from electronic questionnaire asking “How would you describe your usual walking pace?” and was available in 335288 samples. The handgrip strength, available in 335,061 samples, was measured single time for each hand and we used the average value.^12^ 4) Lastly, we included the objective data for physical activity, collected from wrist-band accelerometry.^22^ We used the time for physical activity with moderate-to-vigorous intensity (activity as greater than 100 milligravities in intensity) and the data was available in 71,433 samples.^14^

To calculate the association between genetic instruments and outcome phenotype, linear regression analysis was performed for the SNPs by PLINK 2.0 and the genetic effect sizes were adjusted for age, sex, age×sex, age^2^, and 10 genetic principal components.^23^

### Summary-level MR analysis

The multiplicative random-effects inverse variance-weighted method was the main MR analysis following the current guideline. The method allows balanced pleiotropic effects of the utilized variants.^19,24^ Next, the weighted median method, which relaxes the MR assumptions for up to half of instruments, was used.^18^ MR–Egger regression with bootstrapped standard error was performed.^17^ MR–Egger regression is a method that provides pleiotropy-robust causal estimates under the attainment of the InSIDE assumption. Another strength of the method is that the MR-Egger intercept can be calculated, a measure to assess the presence of a directional pleiotropy. If the intercept is significantly non-zero (P < 0.05), a presence of a directional pleiotropy is suspected. However, MR-Egger regression suffers from weak statistical power when compared to other MR methods. The summary-level MR analysis was performed using the “TwoSampleMR” package in R (version 0.4.26).^25^ As we are performing analysis towards five outcome traits, Bonferroni-adjusted two-sided P value of < 0.05/5 by the inverse variance-weighted method was used to indicate statistical significance. The causal estimates were scaled to the effects from two-fold increase in prevalence of being a smoker by multiplying 0.693 to the raw causal estimates.^26^

### Power calculation for MR analysis

Post-hoc power calculation for MR analysis is available in an online tool (URL: https://sb452.shinyapps.io/power/). We followed the method suggested by S.Burgess et al. (URL: http://mendelianrandomization.com/) to calculate the power when genetically predicting binary exposure.^26,27^ We calculated beta^2^×2×MAF×(1-MAF), where MAF indicates minor allele frequency, of each genetic instruments and the sum of the values was the coefficient of determination of exposure. Despite the limitation, this approach of using odds ratios as a proxy for prevalence ratio was necessary as genetic beta effect sizes were not scaled to prevalence increase of a unit of an exposure in GWAS. The effect sizes of causal estimates by the inverse variance–weighted method towards outcome traits was used as the effect size of the causal effect to calculate the statistical power of MR analysis.

## Results

### Characteristics of the outcome data

The final study sample included as the outcome data for MR analysis has median age of 58 years old [interquartile range 51; 63] (Table 1). Male sex was identified for 46.3%, median body mass index was 26.7 [interquartile range 24.1; 29.9] kg/m^2^. The prevalence of diabetes mellitus was 4.8% and total of 20.9% and 17.5% of the samples were taking hypertension and dyslipidemia medications, respectively.

**Table 1.** Summary-level MR results.

| **Variables** | **Study sample**<br>**(N = 337138)** |
| --- | --- |
| Age (years) | 58.00 [51.00;63.00] |
| Sex |  |
| Female | 181027 (53.7%) |
| Male | 156111 (46.3%) |
| Hypertension medication users | 70021 (20.9%) |
| Dyslipidemia medication users | 58533 (17.5%) |
| Diabetes mellitus | 16179 (4.8%) |
| Systolic BP | 136.5 [125.0;149.5] |
| Diastolic BP | 82.0 [75.5;89.0] |
| Hb A1c (mmol/L) | 35.1 [32.7;37.7] |
| Body mass index (kg/m<sup>2</sup>) | 26.72 [24.13;29.85] |
| Waist circumference (cm) | 90 [80;99] |
| Smoking history |  |
| None-smoker | 183640 (54.66%) |
| Ex-smoker | 118403 (35.24%) |
| Current-smoker | 33922 (10.10%) |
| **Outcomes** |  |
| Telomere (standardized Z-score) | -0.03 [-0.67;0.61] |
| Appendicular lean mass index | 0.85 [0.72;1.02] |
| Handgrip strength (average of measurements for each hand) | 29.00 [22.50;39.00] |
| Walking pace |  |
| Slow | 25105 (7.5%) |
| Average | 175921 (52.5%) |
| Brisk | 134262 (40.0%) |
| Accelerometry-determined moderate-to-vigorous physical activity (hours/week) | 11.09 [7.56;14.95] |

Categorical variables are presented as N (%) and continuous variables as median (interquartile ranges).

The median appendicular lean mass index was 0.85 [interquartile range 0.72; 1.02] and handgrip strength was 29.0 [interquartile range 22.5; 39.0] kg, respectively. There were 40.0% individuals reported brisk walking pace, while 52.5% reported average and 7.5% reported slow walking pace, respectively. The median time for moderate-to-vigorous physical activity, measured by wristband accelerometry, was 11.09 [interquartile range 7.56; 14.95] hours per week.

### Genetic instruments

Total 377/378 SNPs were identified in the UK Biobank data (Supplementary Table 1). Those without genome-wide significant association with an outcome trait were included as the genetic instruments in the according analysis. In addition, there were 14 SNPs with genome-wide significant association with other phenotypes such as education attainment, inflammatory bowel disease, malignancy, and body mass index, identified from the GWAS catalog. Thus, the SNPs were disregarded in the additional sensitivity analysis.

### Summary-level MR analysis results

Genetically predicted being a regular smoker was significantly associated with shorter telomere length [-0.041 (-0.051, -0.031), P < 0.001] (Figure 2 and Table 2). The results remained significant by MR-Egger regression with bootstrapped standard error [-0.027 (-0.059, 0.000), P = 0.048] and weighted median method [-0.043 (-0.059, -0.028), P < 0.001]. Also, the MR-Egger regression intercept P value indicated non-significant pleiotropic effect (P = 0.65), supporting the main causal estimates. The results were similarly replicated even after disregarding the SNPs reported to be associated with other traits.

**Figure 1.**
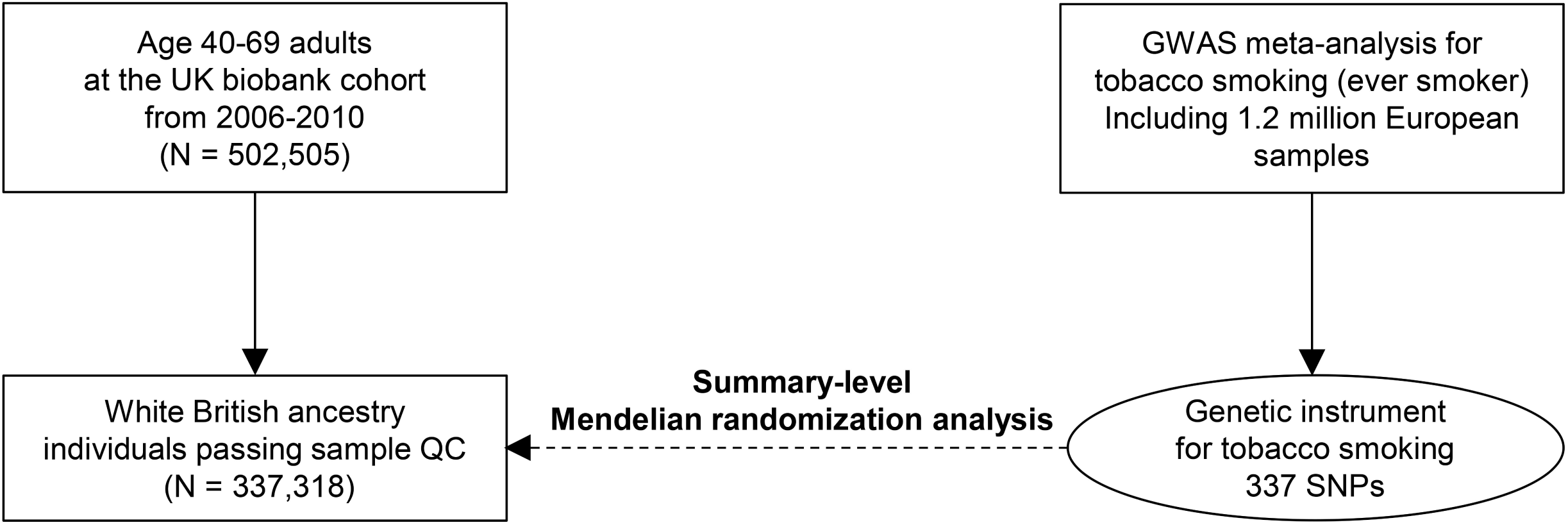
Study flow diagram. GWAS = genome-wide association study, QC = quality control, SNPs = single nucleotide polymorphism

**Figure 2.**
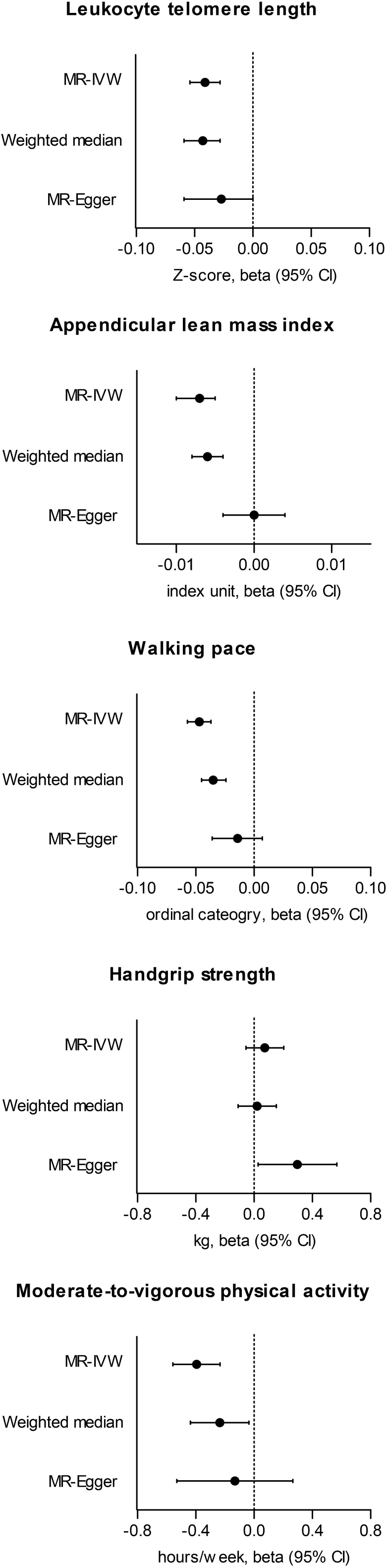
Causal estimates from the Mendelian randomization analysis. The x-axes show the causal estimates. The esimates by MR-IVW (multiplicative random-effect inverse variance weighted method), weighted median, and MR-Egger regression with bootstrapped standard error are presented. The outcome phenotypes were leukocyte telomere length (log-transoformed, standardized to Z-score), appendicular lean mass index (appendicular lean mass/body mass index), walking pace (self-reported ordinal category, slow, average, and brisk), handgrip strength (kg, average from two measurements from each hand), and wristband accelerometer defined time for moderate-to-vigorous physical activity. Causal estimates are scaled towards 2-fold increase in prevalence of tobacco smoking.

**Table 2.**
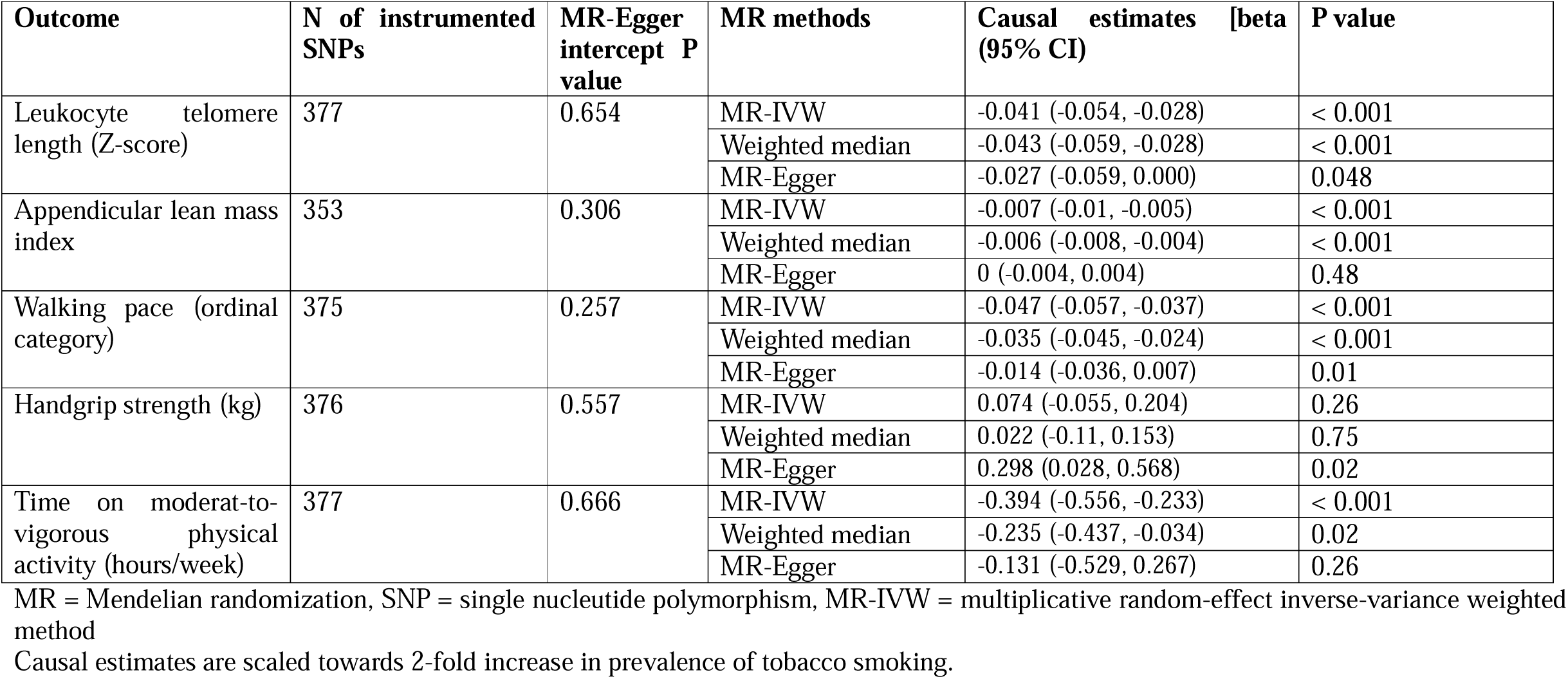
Causal estimates by summary-level MR analysis.

Genetically predicted being a regular smoker was also significantly associated with lower appendicular lean mass index [-0.007 (-0.010, -0.005), P < 0.001]. Again, the results remained significant by weighted median method [-0.006 (-0.008, -0.004), P < 0.001]. Although the MR-Egger regression intercept was approximately zero (P = 0.31), the causal estimates by the MR-Egger regression was attenuated [0.000 (-0.004, 0.004), P = 0.48]. Yet, the main causal estimate remained significant in the sensitivity analysis disregarding SNPs associated with potential confounders.

The causal estimates towards walking pace indicated a higher genetic liability for being a smoker was significantly associated with having a slower walking pace category [-0.047 (-0.057, -0.037), P < 0.001]. The results were similar by the weighted median methods [-0.035 (-0.045, -0.024), P < 0.001] and was marginal for MR-Egger regression [-0.014 (-0.036, 0.007), P = 0.10]. The MR-Egger regression intercept P value indicated low probability of directional pleiotropy (P = 0.26) and the results remained similar in the sensitivity analysis with fewer instrumented SNPs. However, the main causal estimates from genetically predicted tobacco smoking towards handgrip strength was nonsignificant [0.074 (-0.055, 0.204), P = 0.26].

The summary-level MR analysis demonstrated that genetically predicted tobacco smoking was significantly associated with lower time spent on moderate-to-vigorous physical activity [-0.394 (-0.556, -0.233), P < 0.001]. The results by the weighted median method [-0.235 (-0.437, -0.034), P = 0.02] and although the results by MR-Egger regression was marginal [-0.131 (-0.529, 0.267), P = 0.26], the direction of the effect size was same as the main estimate and the MR-Egger intercept P value was non-significant (P = 0.67), supporting that the main estimate was less likely to be affected from directional pleiotropy.

### Post-hos power calculation results

The calculated coefficient for determination was 0.055 except for analysis with the appendicular lean mass index outcome, which yielded 0.051 coefficient, as large number of SNPs were disregarded due to their genome-wide significant association with the outcome itself (Table 3). The post-hoc power towards the outcome phenotypes were sufficient for telomere length (99.8%), appendicular lean mass (100%), walking pace (100%), and accelerometer-defined moderate-to-vigorous physical activity (100%). However, the relative effect sizes of the main causal estimate towards handgrip strength considering the standard deviation was relatively small compared to the others, and the power was limited (10.5%) in the analysis with the handgrip strength outcome.

**Table 3.** Post-hoc power calculation for MR analysis.

| Outcome trait | Outcome sample size | N of instrumented SNPs | Coefficient of determination <sup>a</sup> | Causal estimate effect size (standard deviation change) <sup>b</sup> | Power (%) <sup>c</sup> |
| --- | --- | --- | --- | --- | --- |
| Telomere length | 326075 | 377 | 0.055 | 0.041 | 99.8 |
| Appendicular lean mass index | 329903 | 353 | 0.051 | 0.059 | 100 |
| Walking pace | 335288 | 375 | 0.055 | 0.111 | 100 |
| Handgrip strength | 335061 | 376 | 0.055 | 0.010 | 10.5 |
| Time for moderate-to-vigorous physical activity | 71443 | 377 | 0.055 | 0.097 | 100 |

SNP = single nucleotide polymorphism
<sup>a</sup> Coefficient of determination was calculated by summing the $\beta^2 \times 2 \times \text{MAF} \times (1 - \text{MAF})$ , where MAF indicates minor allele frequency, of each genetic instruments. The coefficient is a proxy value as the current MR analysis tested a binary tobacco smoking exposure, different from a conventional analysis testing the effects from continuous exposure.
<sup>b</sup> Standardized causal estimate effect sizes were driven from the main causal estimates by the inverse-variance weighted method, divided by the standard deviation of the outcome phenotype.
<sup>c</sup> Statistical power was calculated from an online power calculator (URL: https://sb452.shinyapps.io/power/), and we followed the method suggested by S.Burgess et al. (URL: http://mendelianrandomization.com/) to calculate the power in MR analysis with binary exposure.

## Discussion

In this MR analysis, we identified that genetically predicted tobacco smoking was significantly associated with shorter leukocyte telomere length, lower appendicular lean mass index, slower walking pace, and lower physical activity. We considered important aspects of MR analysis to demonstrate causal estimates, and the results is supportive for that tobacco smoking may be a causal risk factor for ageing traits. The study highlights reducing tobacco use may be helpful to reduce the rapidly increasing burden of ageing epidemic in modern medicine.

Ageing traits are important medical dimensions related to various complications (e.g. cardiovascular disease, metabolic syndrome) and mortality.^4,5^ The velocity of ageing differs among individuals, and those with accelerated ageing suffers from adverse outcomes related to frailty. On the other hand, the causal effect from tobacco smoking, a harmful lifestyle behavior which global awareness has been risen for,^7^ on ageing traits has been yet determined in a population-scale despite the numerous evidence that tobacco exposure accelerates cellular senescence.^8^ In addition, as the association between tobacco smoking and ageing traits occurring in later period of life is confounded by many related traits, conventional observational study may have difficulty revealing the causal linkage because of the confounding effects and issue related to reverse causation.^10^ A previous MR study investigating the adverse effects from tobacco smoking reported null finding towards telomere length shortening,^28^ thus, an additional study with robust genetic instruments and a larger-scale outcome data, ensuring the statistical power, was warranted. By this study, we showed that genetic predisposition for tobacco smoking is significantly associated with adverse ageing traits, through implementing the genetic variants reported from the recent large-scale GWAS meta-analysis. The main strength of this study is that we assessed certain dimensions of ageing phenomenon and frailty. Based on our results, medical society should pay attention on preventing tobacco use in the general population to reduce the socioeconomic burden of ageing traits.

That tobacco smoking causes cellular senescence in organs where direct exposure occurs (e.g. lung) have been established from various experimental studies.^29^ Tobacco smoking has been reported to be a risk factor for skeletal muscle dysfunction, not only in lung disease patients but also in the general population.^30-32^ Smoking directly induces skeletal muscle dysfunction by impaired oxygen deliveries to mitochondria, which the chronic condition may lead to muscle atrophy and sarcopenia. In our study, genetically predicted tobacco smoking was significantly associated with both objective and functional dimensions of sarcopenia and physical activity, suggesting that the effect would be generally present by tobacco use in the general population. In addition, our study suggests that direct effect on cellular senescence, indicated by telomere length attrition, may also contribute to the ageing phenomenon. Future study may consider testing the effects from reducing or cessation of tobacco use on ageing or frailty, further supporting the efforts to educate smoking cessation even in those already has smoking histories. In addition, additional investigation for the mechanisms of the effect from components of tobacco on cellular senescence may reveal future directions to reverse such effect.

There are certain limitations in this study. First, debates may present regarding the significance of the causal estimates by MR-Egger regression.^19^ Some may advocate the causal estimates by MR-Egger regression is supportive for the main estimates as the directions were generally consistent and MR-Egger intercept indicated low probability of a directional pleiotropy.^33^ However, as tobacco smoking is a complex social behavior, the possibility of a pleiotropic pathway may still needs to be considered. Second, there was certain sample overlap in the two-sample MR as the GWAS meta-analysis for tobacco smoking included the UK Biobank data. If instrumental power is weak, this may cause bias in the causal estimates,^34^ however, considering that most analysis were performed under sufficient power, such issue may not be substantial. Third, the MR statistical power for handgrip strength was not secured. However, as the main reason may be the actual effect size of the causal estimate is small, leaving handgrip strength to have lower priority outcome in consideration for the causal effects from tobacco smoking. Lastly, performing MR analysis with binary exposure tests the association between genetic liability and outcomes.^26^ However, utilizing quantitative traits (e.g. smoking heaviness) as exposure phenotypes was not possible for tobacco smoking as testing the causal estimates only in a subgroup (e.g. smokers) causes collider bias.^35^

In conclusion, tobacco smoking may be a causal factor for telomere shortening, muscle mass loss, slow gait speed, and physical inactivity. Healthcare providers should remind the harmful effects from tobacco smoking, particularly affecting the various dimensions of ageing traits.

## Data Availability

The data used for this study is available in the public database. The CKDGen data is openly available in the consortium website (URL: https://ckdgen.imbi.uni-freiburg.de/) and the UK Biobank data is accessible after acquiring approval from the organization (application No. 53799).

## Authors’ contribution

The corresponding author attests that all listed authors meet the authorship criteria and that no others meeting the criteria have been omitted. SP, HL, KSK, KWJ, and DKK contributed to the conception and design of the study. SL, SK, YK, YCK, SSH, JPL, KWJ, CSL, YSK, and DKK provided statistical advice and interpreted the data. SP, KSK performed the main statistical analysis, assisted by SK, SL and YK. HL, JPL, KWJ, CSL, YSK, and DKK provided advice regarding the data interpretation. YCK, SSH, HL, JPL, KWJ, CSL, and YSK provided material support during the study. SP and DKK had full access to all data in the study and take responsibility for the integrity of the data and the accuracy of the data analysis. All authors participated in drafting the manuscript. All authors reviewed the manuscript and approved the final version to be published.

## Acknowledgements

The study was based on the data provided by the UK Biobank consortium (application No. 53799). We thank the investigators of the CKDGen consortium who provided the summary statistics for the outcome data of this study.

## References

1. Partridge L, Deelen J, Slagboom PE. Facing up to the global challenges of ageing. Nature 561:45–56, 2018

2. Beard JR, Officer A, de Carvalho IA, Sadana R, Pot AM, Michel JP, et al. The World report on ageing and health: a policy framework for healthy ageing. Lancet 387:2145–2154, 2016

3. Cruz-Jentoft AJ, Baeyens JP, Bauer JM, Boirie Y, Cederholm T, Landi F, et al. Sarcopenia: European consensus on definition and diagnosis: Report of the European Working Group on Sarcopenia in Older People. Age Ageing 39:412–423, 2010

4. Studenski S, Perera S, Patel K, Rosano C, Faulkner K, Inzitari M, et al. Gait speed and survival in older adults. Jama 305:50–58, 2011

5. Leong DP, Teo KK, Rangarajan S, Lopez-Jaramillo P, Avezum A, Jr., Orlandini A, et al. Prognostic value of grip strength: findings from the Prospective Urban Rural Epidemiology (PURE) study. Lancet 386:266–273, 2015

6. Wang Q, Zhan Y, Pedersen NL, Fang F, Hägg S. Telomere Length and All-Cause Mortality: A Meta-analysis. Ageing Res Rev 48:11–20, 2018

7. Ghebreyesus TA. Progress in beating the tobacco epidemic. Lancet 394:548–549, 2019

8. Nyunoya T, Monick MM, Klingelhutz A, Yarovinsky TO, Cagley JR, Hunninghake GW. Cigarette smoke induces cellular senescence. Am J Respir Cell Mol Biol 35:681–688, 2006

9. Astuti Y, Wardhana A, Watkins J, Wulaningsih W. Cigarette smoking and telomere length: A systematic review of 84 studies and meta-analysis. Environ Res 158:480–489, 2017

10. Davies NM, Holmes MV, Davey Smith G. Reading Mendelian randomisation studies: a guide, glossary, and checklist for clinicians. Bmj 362:k601, 2018

11. Bycroft C, Freeman C, Petkova D, Band G, Elliott LT, Sharp K, et al. The UK Biobank resource with deep phenotyping and genomic data. Nature 562:203–209, 2018

12. Park S, Lee S, Kim Y, Lee Y, Kang MW, Kim K, et al. Relation of Poor Handgrip Strength or Slow Walking Pace to Risk of Myocardial Infarction and Fatality. Am J Cardiol 162:58–65, 2022

13. Park S, Lee S, Kim Y, Cho S, Kim K, Kim YC, et al. A Mendelian randomization study found causal linkage between telomere attrition and chronic kidney disease. Kidney Int 100:1063–1070, 2021

14. Park S, Lee S, Kim Y, Lee Y, Kang MW, Kim K, et al. Causal effects of physical activity or sedentary behaviors on kidney function: an integrated population-scale observational analysis and Mendelian randomization study. Nephrol Dial Transplant 37:1059–1068, 2022

15. Sudlow C, Gallacher J, Allen N, Beral V, Burton P, Danesh J, et al. UK biobank: an open access resource for identifying the causes of a wide range of complex diseases of middle and old age. PLoS Med 12:e1001779, 2015

16. Liu M, Jiang Y, Wedow R, Li Y, Brazel DM, Chen F, et al. Association studies of up to 1.2 million individuals yield new insights into the genetic etiology of tobacco and alcohol use. Nat Genet 51:237–244, 2019

17. Bowden J, Davey Smith G, Burgess S. Mendelian randomization with invalid instruments: effect estimation and bias detection through Egger regression. Int J Epidemiol 44:512–525, 2015

18. Bowden J, Davey Smith G, Haycock PC, Burgess S. Consistent Estimation in Mendelian Randomization with Some Invalid Instruments Using a Weighted Median Estimator. Genet Epidemiol 40:304–314, 2016

19. Burgess S, Davey Smith G, Davies NM, Dudbridge F, Gill D, Glymour MM, et al. Guidelines for performing Mendelian randomization investigations. Wellcome Open Res 4:186, 2019

20. Park S, Lee S, Kim Y, Lee Y, Kang MW, Kim K, et al. Atrial fibrillation and kidney function: a bidirectional Mendelian randomization study. Eur Heart J 42:2816–2823, 2021

21. Studenski SA, Peters KW, Alley DE, Cawthon PM, McLean RR, Harris TB, et al. The FNIH sarcopenia project: rationale, study description, conference recommendations, and final estimates. J Gerontol A Biol Sci Med Sci 69:547–558, 2014

22. Doherty A, Jackson D, Hammerla N, Plötz T, Olivier P, Granat MH, et al. Large Scale Population Assessment of Physical Activity Using Wrist Worn Accelerometers: The UK Biobank Study. PLoS One 12:e0169649, 2017

23. Chang CC, Chow CC, Tellier LC, Vattikuti S, Purcell SM, Lee JJ. Second-generation PLINK: rising to the challenge of larger and richer datasets. Gigascience 4:7, 2015

24. Skrivankova VW, Richmond RC, Woolf BAR, Davies NM, Swanson SA, VanderWeele TJ, et al. Strengthening the reporting of observational studies in epidemiology using mendelian randomisation (STROBE-MR): explanation and elaboration. Bmj 375:n2233, 2021

25. Hemani G, Zheng J, Elsworth B, Wade KH, Haberland V, Baird D, et al. The MR-Base platform supports systematic causal inference across the human phenome. Elife 7, 2018

26. Burgess S, Labrecque JA. Mendelian randomization with a binary exposure variable: interpretation and presentation of causal estimates. Eur J Epidemiol 33:947–952, 2018

27. Zeng H, Ge J, Xu W, Ma H, Chen L, Xia M, et al. Type 2 Diabetes Is Causally Associated With Reduced Serum Osteocalcin: A Genomewide Association and Mendelian Randomization Study. J Bone Miner Res, 2021

28. Rode L, Bojesen SE, Weischer M, Nordestgaard BG. High tobacco consumption is causally associated with increased all-cause mortality in a general population sample of 55,568 individuals, but not with short telomeres: a Mendelian randomization study. Int J Epidemiol 43:1473–1483, 2014

29. University of Essex. Institute for Social and Economic Research, NatCen Social Research, Kantar Public. (2016). Understanding Society: Waves 1-6, 2009-2015. In: Service UD, ed. 8th Edition. ed.

30. Chan SMH, Cerni C, Passey S, Seow HJ, Bernardo I, van der Poel C, et al. Cigarette Smoking Exacerbates Skeletal Muscle Injury without Compromising Its Regenerative Capacity. Am J Respir Cell Mol Biol 62:217–230, 2020

31. Castillo EM, Goodman-Gruen D, Kritz-Silverstein D, Morton DJ, Wingard DL, Barrett-Connor E. Sarcopenia in elderly men and women: the Rancho Bernardo study. Am J Prev Med 25:226–231, 2003

32. Locquet M, Bruyère O, Lengelé L, Reginster JY, Beaudart C. Relationship between smoking and the incidence of sarcopenia: The SarcoPhAge cohort. Public Health 193:101–108, 2021

33. Burgess S, Thompson SG. Interpreting findings from Mendelian randomization using the MR-Egger method. Eur J Epidemiol 32:377–389, 2017

34. Burgess S, Davies NM, Thompson SG. Bias due to participant overlap in two-sample Mendelian randomization. Genet Epidemiol 40:597–608, 2016

35. Gkatzionis A, Burgess S. Contextualizing selection bias in Mendelian randomization: how bad is it likely to be? Int J Epidemiol 48:691–701, 2019

