## Supplementary Material for "Causal linkage of tobacco smoking with ageing traits: a Mendelian randomization analysis towards telomere attrition and frailty"

**Online supplements**

Title

S Park et al.

**Table of contents**

**Supplemental Table 1**. Information of genetic instruments and their association with outcome phenotypes.

**Supplemental Table 2.** Causal estimates by summary-level MR analysis after disregarding SNPs associated with other phenotypes by the GWAS catalog screening.

**Supplemental Table 2.** Causal estimates by summary-level MR analysis after disregarding SNPs associated with other phenotypes by the GWAS catalog screening.

| **Outcome** | **N of instrumented SNPs** | **MR-Egger intercept P value** | **MR methods** | **Causal estimates [beta (95% CI)** | **P value** |
| --- | --- | --- | --- | --- | --- |
| Leukocyte telomere length (Z-score) | 363 | 0.530 | MR-IVW | -0.04 (-0.053, -0.027) | < 0.001 |
|  |  |  | Weighted median | -0.041 (-0.057, -0.025) | < 0.001 |
|  |  |  | MR-Egger | -0.025 (-0.058, 0.007) | 0.07 |
| Appendicular lean mass index | 344 | 0.356 | MR-IVW | -0.007 (-0.008, -0.006) | < 0.001 |
|  |  |  | Weighted median | -0.005 (-0.007, -0.003) | < 0.001 |
|  |  |  | MR-Egger | 0 (-0.004, 0.004) | 0.47 |
| Walking pace (ordinal category) | 362 | 0.209 | MR-IVW | -0.046 (-0.056, -0.035) | < 0.001 |
|  |  |  | Weighted median | -0.034 (-0.045, -0.024) | < 0.001 |
|  |  |  | MR-Egger | -0.015 (-0.037, 0.007) | 0.09 |
| Handgrip strength (kg) | 362 | 0.463 | MR-IVW | 0.125 (-0.006, 0.256) | 0.06 |
|  |  |  | Weighted median | 0.072 (-0.057, 0.201) | 0.28 |
|  |  |  | MR-Egger | 0.38 (0.125, 0.635) | 0.001 |
| Time on moderat-to-vigorous physical activity (hours/week) | 363 | 0.542 | MR-IVW | -0.376 (-0.538, -0.214) | < 0.001 |
|  |  |  | Weighted median | -0.235 (-0.44, -0.031) | 0.02 |
|  |  |  | MR-Egger | -0.034 (-0.427, 0.359) | 0.44 |

MR = Mendelian randomization, SNP = single nucleutide polymorphism, MR-IVW = multiplicative random-effect inverse-variance weighted method

Causal estimates are scaled towards 2-fold increase in prevalence of tobacco smoking.
